# Myocardial Stiffness Tracking & Assessment Toolkit Driven by Artificial Intelligence For Reproducible Benchmarking of Temporal Segmentation and Shear-Wave Velocity Stabilization on Synthetic Data

**DOI:** 10.64898/2026.09.17.26363353

**Authors:** Terry Tsebro, Abdullah Safi, Bangcheng Wang, Luca Hutchinson, Mark Noge, Kamil Chaudhry, Laith Alzoubi, Austin Hua, Nimish Ray, Parham Baghbanbashi, Antonin Chianale, Meryem Karayunusoglu, Yimin Xiu, Amar Saed, Wagih Ghobriel, Aimen Malik, Dennis D. Fernandes

**Author notes:** Corresponding authors: Dennis D. Fernandes, Aimen Malik. Current mailing address (Yimin Xiu, Aimen Malik, Dennis D. Fernandes).

## Abstract

**Propose:** Routine echocardiographic assessment can be affected by operator variability and time-consuming post-processing, which may limit rapid quantitative analysis. Real-time cine ultrasound demands low-latency automated processing for clinical usage. Existing approaches lack standardized evaluation frameworks, verified edge deployment, and structured reproducibility guarantees. To address these limitations, we built Myocardial Stiffness Tracking & Assessment Toolkit driven by Artificial Intelligence (MyoSTAT.AI). We report a deterministic, fully reproducible pipeline that performs real-time segmentation and SWE velocity estimation, evaluated entirely on synthetic data.

**Approach:** Four U-Net-based architectures spanning single-frame and temporal designs were evaluated via systematic ablation: a 2D baseline, a 2.5D stacked-frame model, a 3D volumetric model, and a ConvLSTM variant. Experiments used synthetic reference corpora with split accounting the segmentation corpus contained 1,200 synthetic frames, and the SWE corpus contained 900 synthetic velocity-field cases. The SWE branch used Radon-transform-based propagation-direction estimation, time-domain shear-wave speed estimation, and configurable temporal stabilization on synthetic velocity fields. TensorRT FP16 deployment benchmarks were performed separately for the exported UNet2.5D segmentation model on RTX 3060 and Jetson Orin Nano hardware.

**Results:** On the synthetic evaluation quantities, the ConvLSTM variant achieved the highest segmentation accuracy (Dice 0.994, IoU 0.987), an 18.6% improvement over the 2D baseline (Dice 0.808). A 2.5D model with a 3-frame temporal window achieved Dice 0.983 at substantially lower latency (433 ms vs. 1193 ms). TensorRT FP16 deployment yielded 341 FPS on the RTX 3060 and 90.4 FPS on the Jetson Orin Nano. Penalized least-squares temporal stabilization (smoothn, λ=10) reduced shear-wave speed CoV by 63.8% at 2.7 ms latency overhead per frame, with no loss of edge preservation.

**Conclusions:** We demonstrate a deterministic, reproducible computational benchmarking framework for cardiac segmentation and SWE velocity-field stabilization, evaluated entirely on synthetic data, together with preliminary inference feasibility on workstation and embedded hardware. These results represent a synthetic proof-of-concept rather than a validated clinical or operator-independent tool; acquisition of real echocardiographic data and clinical validation are required before any claim of diagnostic or deployment readiness can be made.

## Introduction

Echocardiographic cine imaging is widely used to assess cardiac anatomy and motion, but frame-by-frame segmentation can be time-consuming and operator-dependent. In this study, the segmentation task is defined as binary target-region mask prediction from synthetic cardiac ultrasound cine frames, where temporal models use one or more adjacent grayscale frames to predict the target-frame mask. Accordingly, the present study evaluates temporal segmentation and inference feasibility as a computational benchmark rather than as a clinical diagnostic system or validated treatment decision support. AI-assisted echocardiographic segmentation is increasingly evaluated using clinically annotated datasets and video-based benchmarks rather than generic ultrasound examples alone. CAMUS provides annotated two- and four-chamber 2D echocardiographic sequences for multi-structure segmentation (Leclerc et al., 2019), and EchoNet-Dynamic provides large-scale apical four-chamber videos for left-ventricular tracing and functional assessment (Ouyang et al., 2020). Beyond framewise U-Net segmentation, recent work has examined temporal consistency across the cardiac cycle (Painchaud et al., 2022), while real-time echocardiographic pipelines connect segmentation to downstream measurements and deployment constraints (Smistad et al., 2020). The present study is therefore positioned as a synthetic-domain benchmark of temporal U-Net-based segmentation and hardware-specific inference feasibility, complementary to clinical-dataset validation rather than a substitute for current solutions.

Temporal modeling in echocardiography should be distinguished from training paradigms such as supervised or self-supervised learning. Architectures that use temporal information can include recurrent units, attention-based modules, optical-flow tracking, and temporal-consistency constraints. Lucas-Kanade optical flow is a classical motion-tracking method rather than a recent deep temporal architecture. Recent echocardiographic temporal-consistency work has specifically targeted frame-to-frame stability across the cardiac cycle (Painchaud et al., 2022). In this manuscript, the temporal U-Net variants are compared as supervised segmentation architectures, as opposed to self-supervised learning methods for benchmarking purposes as applied to the assessment of myocardial stiffness, a key biological indicator of reduced elasticity of the myocardium, the muscle that plays a key role in cardiac function.

Here, we focus on shear wave elastography (SWE) as a model metric which estimates tissue stiffness from shear-wave propagation speed, which rises with stiffness. Cardiac shear waves may arise from natural mechanical events, such as valve closure or from an acoustic radiation force push, and the two paradigms differ in excitation, required frame rate, and data type. SWE has been applied to myocardial stiffness and strain assessment (Correia et al., 2017). The present work operates on synthetic shear-wave velocity fields and estimates shear-wave speed; the excitation source and acquisition physics are not modeled here and are required for real-data validation. However, existing limitations of these approaches include the need for real-time processing, data acquisition challenges, and issues with noise robustness. There are still significant research gaps in this area, particularly with regard to improving SWE methods for clinical use. Studies have focused on developing more robust and efficient algorithms, including those using convolutional neural networks (CNNs) and recurrent neural networks (RNNs). However, further work is needed to address the challenges of real-time processing, data acquisition, and noise robustness.

The deployment of AI-assisted image analysis tools for cardiac ultrasound also presents several concerns. Real-time inference imposes concrete constraints: per-frame end-to-end latency low enough to keep pace with the acquisition frame rate, a memory footprint within the device budget and, on battery-powered edge hardware, bounded power draw. These tools must integrate seamlessly into existing clinical workflows, which demand a high degree of flexibility and adaptability. Device constraints such as limited processing power and memory also require careful consideration.

Furthermore, hardware integration and deployment feasibility of these programs can pose significant challenges, as studies suggest that deploying AI models to production environments requires careful consideration of model interpretability, explainability, and robustness. This motivates efficient models suitable for embedded deployment. The Myocardial Stiffness Tracking & Assessment Toolkit driven by Artificial Intelligence (MyoSTAT.AI) is a deterministic, reproducible benchmarking framework for temporal echocardiographic segmentation and shear-wave velocity-field stabilization, evaluated entirely on synthetic data. The contributions of this work, separate into both scientific findings and reproducibility tooling, where we characterize five segmentation design axes (architecture, temporal window, loss function, augmentation, and dropout) through a single-variable ablation, quantifying their effect on Dice, IoU and latency under a fixed synthetic regime. Further, we catalog seven engineering failure modes, each with quantitative evidence, candidate root causes, and mitigations. As such, we provide a deterministic temporal data loader and synthetic data generator with fixed-seed guarantees. This allowed for the implementation of a configurable shear-wave velocity-field stabilization pipeline whose smoothing presets improve the temporal consistency of the estimated velocity fields, a step downstream of and independent from segmentation. Here, we release a version of benchmark harness with reference-dataset registration and integrity checks, capable of the generation of evidence bundles that package configurations, per-variant metrics, and checksums for independent verification.

## Materials & Methods

### Pipeline & Architecture Family

We evaluate four segmentation architectures spanning the 2D-to-temporal design spectrum. All architectures use a U-Net-style encoder-decoder topology with skip connections, where the input dimensionality and normalization layers differ. UNet2D processes a single greyscale frame of shape (B, 1, H, W) and produces a single logit map of shape (B, 1, H, W). It serves as the 2D reference baseline with 7.696193 M trainable parameters. UNet2.5D receives a tensor of shape (B, N, H, W), with N consecutive frames stacked as input channels. The architecture comparison uses N = 3, while N ∈ {1, 3, 5, 7} is evaluated in the temporal-window ablation. It produces one logit map of shape (B, 1, H, W), supervised against the center-frame mask, and contains 7.702977 M trainable parameters when N = 3. UNet3D receives a tensor of shape (B, N,) and, reshapes it internally to (B, 1, N, H, W), and applies volumetric convolutions jointly across the spatial and temporal dimensions. The architecture comparison uses N = 6, and the decoded temporal features are reduced to one center-frame logit map of shape (B, 1, H, W). UNet+ConvLSTM receives a five-frame tensor of shape (B, 5, 1, H, W), encodes each frame with a shared U-Net encoder, and aggregates the bottleneck features using two ConvLSTM layers with 64 hidden channels per layer and 3 × 3 recurrent kernels (Shi et al., 2015). In the architecture ablation, it uses sequence-to-one inference and decodes the final time step to one logit map of shape (B, 1, H, W). No dropout is applied inside the ConvLSTM recurrent cells; Dropout2d with p = 0.1 is applied only in the surrounding convolutional blocks. The appropriate activation is applied inside each loss during training (i.e., a sigmoid before soft Dice, or binary cross-entropy computed directly from logits for the BCE variants), and at inference a sigmoid is applied and the probability is thresholded at 0.5 for binary segmentation. Input spatial resolution is 128×128 for ablation runs and 256×256 for benchmark evaluation.

### Temporal Data Loading

Temporal input sequences are constructed by a custom TemporalDataLoader that retrieves N consecutive frames centered on the target frame, applies configurable stride, and handles boundary conditions via frame repetition. The loader is deterministic given a fixed seed, ensuring identical training and validation batches across runs. Since all current data are synthetic and carry no patient identity, patient-level stratification does not apply; the independence unit is the individual synthetic sample, and each generated sample is assigned to exactly one split so that no sample appears in more than one split. Split manifests are verified against SHA-256 integrity records before any benchmark run. Patient-level stratification is reserved for future clinical data. For ablation runs, all data is synthetic: frames are generated by a reproducible synthetic data generator seeded via determinism.seed = 42 in each variant config. Synthetic masks have a target foreground density of 0.35. For testing, we used several training configurations to test and achieve a set of outcomes. These details of the various training configurations are documented in **Table 1**.

**Table 1.** Segmentation Pipeline Training Configurations.

| Parameter | Ablation runs | Full Training |
| --- | --- | --- |
| Optimizer | Adam | Adam |
| Learning rate | $1 \times 10^{-3}$ | $1 \times 10^{-3}$ |
| Weight decay | $1 \times 10^{-4}$ | $1 \times 10^{-4}$ |
| LR schedule | — | ReduceLROnPlateau<br>(factor 0.5, patience 8, min lr $1 \times 10^{-5}$) |
| Batch size | 4 | 2 |
| Training steps | 20 gradient steps | 70 epochs |
| Early stopping | — | Patience 15, monitored: val_dice |
| Seed | 42 | 42 |
| Hardware | CPU FP32 | NVIDIA RTX 3060 Laptop (5.67 GB),<br>CUDA 12.6 |
| Input resolution | 128 x 128 | 256 x 256 |

**Note:** Loss functions evaluated across the loss axis:
dice: Soft Dice loss with smoothing $\epsilon = 1 \times 10^{-6}$
bce: Binary cross-entropy with logits
dice\_bce: Weighted sum of Dice and BCE (BCE weight 0.5)
dice\_focal: Weighted sum of Dice and focal loss (Lin, 2017), $\gamma = 2.0$
Augmentation evaluated across the augmentation axis:
aug\_none: No augmentation
aug\_flip: Random horizontal flip ( $p = 0.5$ )
aug\_flip\_rotation: Flip + random rotation ( $\pm 10^\circ$ )
aug\_full: Flip + rotation + random brightness ( $\pm 0.2$ ) + random contrast ( $\pm 0.2$ )

### Ablation Study Design & Latency Measurement Protocol

The ablation study follows a single-variable isolation protocol: a fixed baseline configuration is defined, and each subsequent variant changes exactly one variable while holding all others constant. Five design axes are evaluated across 17 variants in the trained-weight study: architecture, temporal window, loss function, augmentation, and dropout. Inference latency is measured as wall-clock time (Python time.perf_counter) per frame, including preprocessing, forward pass, and postprocessing. On CUDA, torch.cuda.synchronize() is called before each timing boundary to ensure device completion. A 20-run warmup is performed before the timed window to exclude JIT compilation and driver initialization overhead. Reported statistics: mean, median, standard deviation, p95, p99. Reported ablation latencies are CPU FP32. Benchmark harness latencies include both CPU FP32 and CUDA FP16 configurations (see materials and methods). As a prospective evaluation criterion, we adopt a real-time inference target of ≥30 FPS (≤33 ms per frame), corresponding to typical 30 Hz echocardiographic cine acquisition. This is an engineering benchmark of model inference latency, not a validated clinical requirement. It excludes acquisition and, for the SWE branch, the ∼0.83 s stabilization-window delay noted in the results section.

### Shear Wave Elastography Pipeline & Signal Processing Chain

Raw input is a 3D velocity field of shape (H, W, T). Processing stages: (1) resampling and amplitude masking (threshold 0.1); (2) bandpass filtering [50-450 Hz] along the time axis; (3) background subtraction (per-pixel temporal mean); (4) frame-to-frame displacement estimation by normalized cross-correlation over 5 x 5-pixel spatial windows applied to each consecutive frame pair. The window step is computed as int(window_size x (1 - overlap)), so the configured 0.5 overlap truncates to a 2-pixel step and a realized overlap of 60%; (5) Radon-transform directional estimation over [−90°, 90°] at 1° resolution. These settings are set by the acquisition rate and expected shear wave frequency content. At a 1000 Hz frame rate, a 5-frame window spans 5 ms. The bandpass-filtered wave energy occupies 50-450Hz (**Table 2**), i.e. periods of 2–20 ms; a 5 ms window which therefore covers approximately a quarter to a full cycle of the expected wave components. This balances the temporal resolution against having enough samples per window for a stable cross-correlation estimate, a longer window would smear fast, high-frequency wave fronts, while a shorter one would not contain enough cycles to correlate reliably. Background subtraction via per-pixel temporal mean removes static/DC tissue clutter that does not oscillate at these frequencies, isolating the propagating wave-induced velocity component that cross-correlation subsequently tracks.

**Table 2.** SWE Parameter Description.

| Parameter | Value | Description |
| --- | --- | --- |
| frame_rate | 1000.0 Hz | Acquisition frame rate |
| sound_speed | 1540.0 m/s | Assumed acoustic propagation speed |
| spatial_resolution | 0.1 mm/px | Pixel pitch |
| temporal_resolution | 0.001 s | Frame interval |
| max_velocity | 10.0 m/s | Velocity clipping threshold |
| min_amplitude | 0.1 | Background mask threshold |
| window_size | 5 px | Cross-correlation window |
| overlap | 0.5 configured (2 px step, 60% realized) | Window overlap fraction |
| filter_cutoff | [50, 450] Hz | Bandpass filter bounds |
| radon_angles | -90° to +90°, step 1° | Directional estimation sweep |
| background_removal | mean | Background subtraction method |
| velocity_method | time_domain | Velocity estimation algorithm |

### Temporal Stabilization

Six stabilization methods are evaluated: no stabilization (baseline), three penalized least-squares variants (smoothn conservative / moderate / aggressive, λ ∈ {5, 10, 20}), and two SNR-adaptive window methods (fine and coarse). Let v and u denote the unstabilized and stabilized fields for one velocity component, respectively, each of size H × W × T. After vectorization into HWT-element vectors, u is obtained by minimizing ||v − u||₂² + λuᵀPu, where λ is the smoothn smoothing parameter and P is an HWT × HWT positive-semidefinite anisotropic roughness penalty operator associated with second-order regularization. The operator P is represented implicitly in the discrete-cosine-transform (DCT) basis. The x- and y-velocity components are processed independently, while all H × W × T samples within each component are smoothed jointly. The implementation sets the axis spacings to (10⁶, 10⁶, 1) for (H, W, T), making spatial regularization negligible relative to temporal regularization. The DCT formulation imposes even-symmetric boundary conditions along the temporal axis. Evaluation metric: temporal Coefficient of Variation (CoV). The velocity estimation and signal processing configuration have been summarized in **Table 2** and **Table 3**.

**Table 3.** Stabilization comparison configuration.

| Parameter | Value | Description |
| --- | --- | --- |
| frame_rate | 30.0 Hz | Frame rate for stabilization evaluation |
| filter_cutoff | [1.0, 10.0] Hz | Bandpass filter bounds |
| filter_order | 3 | Butterworth filter order |
| window_size | 25 frames | Correlation window |
| search_range | 5 pixels | Displacement search range |
| overlap | 0.8 (80%) | Window overlap fraction |
| background_removal | none | Background subtraction disabled |
| apply_directional_filter | true | Shear wave directional filter enabled |

### Benchmark harness design & Dataset Versioning & Integrity

All quantitative results are produced by a reproducible benchmark harness parameterized by versioned baseline config JSON files. Each config specifies model architecture, preprocessing, inference precision, runtime device, and a reference to the dataset manifest, ensuring results are attributable to a specific configuration. Reference datasets are registered in a manifest recording dataset ID, version, split file paths, expected file count, and a SHA-256 checksum of the full file listing. The harness verifies the listing checksum before any run, preventing silent data drift. Generator-parameter-level split isolation is enforced across all model configurations; patient-level isolation applies only once clinical data is added.

### Evidence Bundles & Failure Mode Analysis

After each validated experiment, an evidence bundle is generated: a timestamped, self-contained directory containing all config files, per-variant metrics, aggregated results, and a manifest.json with SHA-256 checksums for every file and the git commit hash at generation time. An evidence bundle lets an independent reviewer inspect the recorded configurations, metrics and checksums for a completed experiment without re-running it, once the bundle is made accessible (see Code, Data, and Materials Availability). A structured failure mode taxonomy documents seven failure modes (FM-001–FM-007) derived from ablation results, covering temporal context regression, training instability, over-regularization, augmentation distribution mismatch, compute latency instability, and system-level variance. Each entry includes quantitative evidence, root cause, severity classification, and mitigation strategies.

### Hardware, Datasets and Splits

Two compute environments are referenced: a workstation used for model training and CUDA inference benchmarks, and the NVIDIA Jetson Orin Nano used for embedded deployment latency measurements. Workstation specifications are recorded automatically in the system block of every benchmark.json produced by the benchmark harness, so **Table 4** reflects a specific reference run. All discussed analysis use synthetic data, with clinical acquisition planned for future work (see discussion and conclusion section).

**Table 4.** Testing Environment Hardware Specifications.

| Role | Component | Specification |
| --- | --- | --- |
| Training and CUDA inference | CPU | 11th Gen Intel Core i7-11370H @ 3.30 GHz (4 physical cores, 8 threads) |
| Training and CUDA inference | RAM | 38.87 GB |
| Training and CUDA inference | GPU | NVIDIA GeForce RTX 3060 Laptop (5.67 GB VRAM) |
| Training and CUDA inference | CUDA | 12.6 (driver 13.0.10) |
| Training and CUDA inference | TensorRT | 10.3.0 |
| Training and CUDA inference | PyTorch | 2.2.0+cu121 |
| Training and CUDA inference | OS | Ubuntu 24.04 |
| Training and CUDA inference | Python | 3.11.9 (conda env) |
| Embedded deployment | Device | NVIDIA Jetson Orin Nano Developer Kit (NVIDIA, 2024a) |
| Embedded deployment | JetPack | 6.2.1 (ARM64) (NVIDIA, 2025) |
| Embedded deployment | TensorRT | 10.3 (matched to workstation export) |
| Embedded deployment | Memory Model | CUDA unified memory (zero-copy on integrated GPU) |
| Planned acquisition device | Probe | Butterfly iQ3+ (handheld POCUS) |

All segmentation and SWE results in this paper are derived from synthetic reference corpora produced by our deterministic synthetic data generator. Patient-level metadata is therefore not applicable for the current results; the registered reference datasets will carry that metadata once populated with clinical data. More information on the dataset split is presented in **Table 5**. The 64-sample ablation corpus is regenerated from global_seed = 42 before each run and supports the rapid mini-train ablation regime. The two reference corpora are registered in benchmarks/datasets_manifest.json with SHA-256 integrity checks against the file listing and are versioned independently of the codebase. For SWE, evaluation also draws on the eight synthetic test conditions defined in the safe-operating-ranges configuration (normal signal, high noise, non-propagating, spatial aliasing, temporal aliasing, boundary effect, strong attenuation, and zero signal), exercising the algorithm under controlled failure-mode inputs.

**Table 5.**
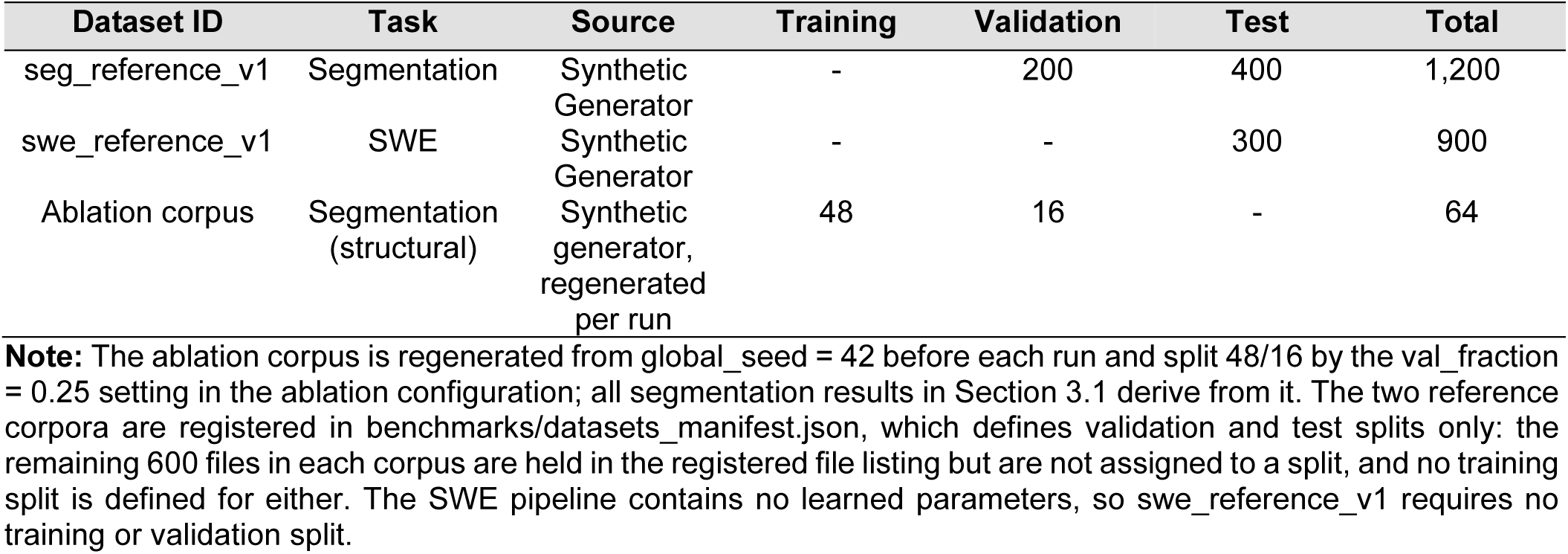
Dataset Split Metrics.

### Evaluation Metrics & Reproducibility Infrastructure

Segmentation accuracy uses Dice and IoU. SWE evaluation uses temporal Coefficient of Variation (CoV) for stability, mean absolute error (MAE) and root-mean-square error (RMSE) for agreement against the synthetic ground-truth shear-wave speeds, and a repeatability CoV across repeated runs. Latency is reported as wall-clock per-frame time using time.perf_counter, with torch.cuda.synchronize calls bracketing all CUDA timings. More information about these table metrics can be found in **Table 6**. Boundary-distance metrics (HD95, ASSD) are out of scope for this work and appear in the future-work plan once clinical data becomes available; surface distance is not meaningful against synthetic ground truth produced by the same generator that defines the masks. Every benchmark and ablation invocation seeds Python hash, NumPy, and PyTorch (CPU and CUDA) from a single source. The benchmark seed bundle in benchmarks/datasets_manifest.json sets global_seed = 1337, python_hash_seed = 0, torch_deterministic_algorithms = true, cudnn_deterministic = true, and cudnn_benchmark = false. The ablation runner uses global_seed = 42 with per-variant seed 42 + variant_index. The two values are kept distinct: 1337 governs benchmark reruns against the fixed reference dataset, and 42 governs the synthetic ablation corpus and its variant ordering.

**Table 6.**
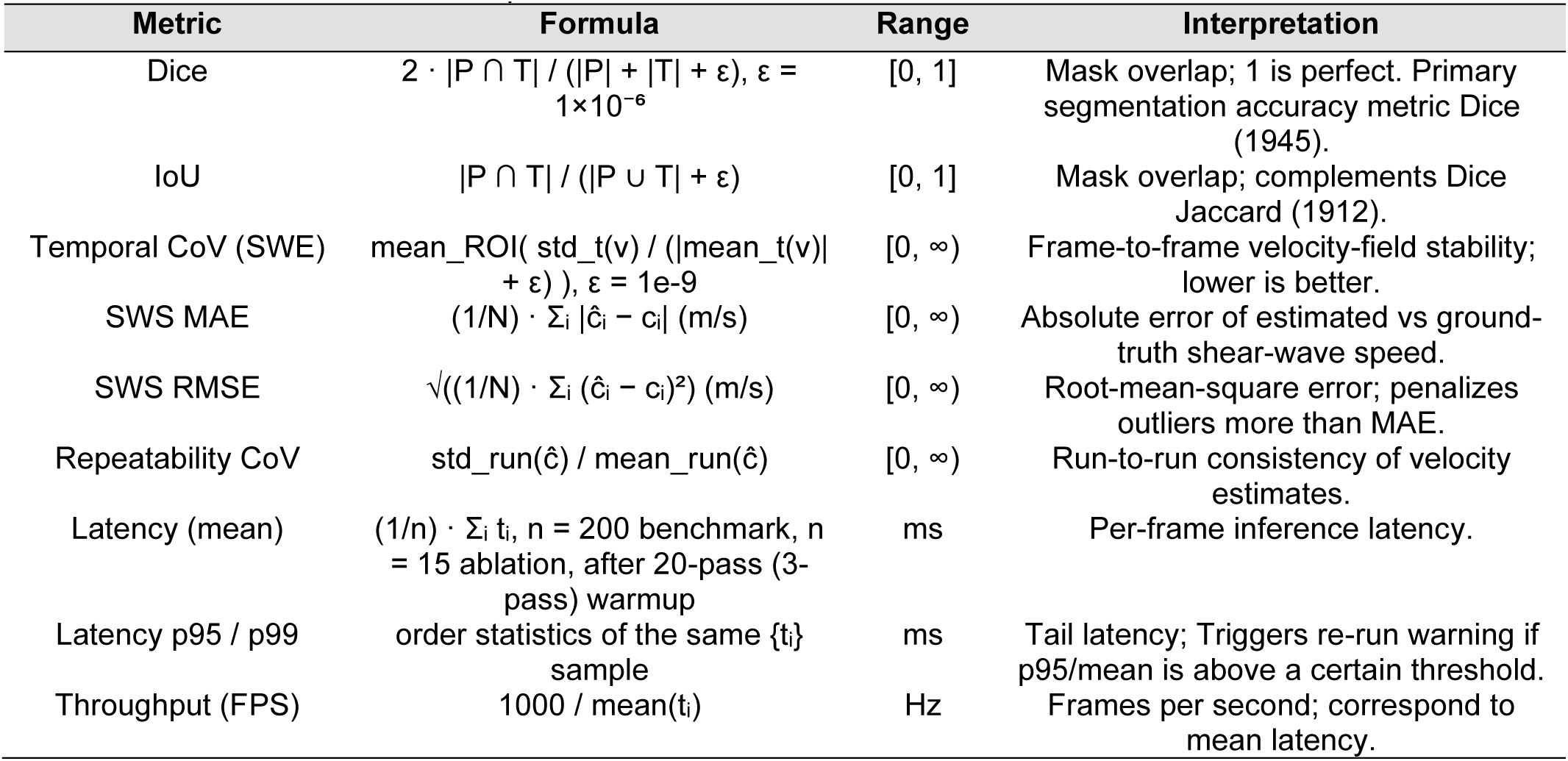
Evaluation of Metrics Description.

Library versions controlling the documentation and toolchain are hard-pinned in docs/requirements.txt to match environment.yml (sphinx==9.0.4, sphinx-rtd-theme==3.1.0, myst-parser==5.0.0, sphinxcontrib-mermaid==2.0.0). The Python runtime is pinned through the internal conda environment (Python 3.11.9). Inference-runtime versions (CUDA, TensorRT, PyTorch) are recorded in each benchmark.json rather than pinned globally, so each reported measurement records the exact stack under which it was captured. The listing checksum described in the methods section covers the file listing rather than per-file contents and remains a placeholder in the manifest until the reference corpora are populated with clinical data; it therefore guards against listing-level drift only and is not yet an active content-integrity check. Because this hash covers the file listing rather than per-file contents and remains a placeholder in the manifest until the reference corpora are populated with clinical data, it currently guards against listing-level drift only and is not yet an active content-integrity check.

## Results

### End-to-end pipeline

Here we summarize the end-to-end pipeline underlying these results, from cine ultrasound acquisition through segmentation, overlay rendering, and SWE velocity estimation to final display (**Figure 1**). The raw cine B-mode frames are resampled and normalized to 256 x 256 before segmentation by the UNet2.5D or ConvLSTM model (see materials and methods), both deployed as FP16 TensorRT engines. The segmentation output feeds two parallel branches: an overlay-render branch that composites the predicted mask onto the source frame, and the SWE branch that applies the Radon-transform velocity estimation and temporal stabilization described in the methods section. Both branches converge in the clinical viewer for display and export. TensorRT FP16 engines compiled from the same exported ONNX model drive both deployment targets, sustaining 341 FPS on the workstation (RTX 3060) and 90.4 FPS on the Jetson Orin Nano via zero-copy unified memory (see materials and methods).

**Figure 1.**
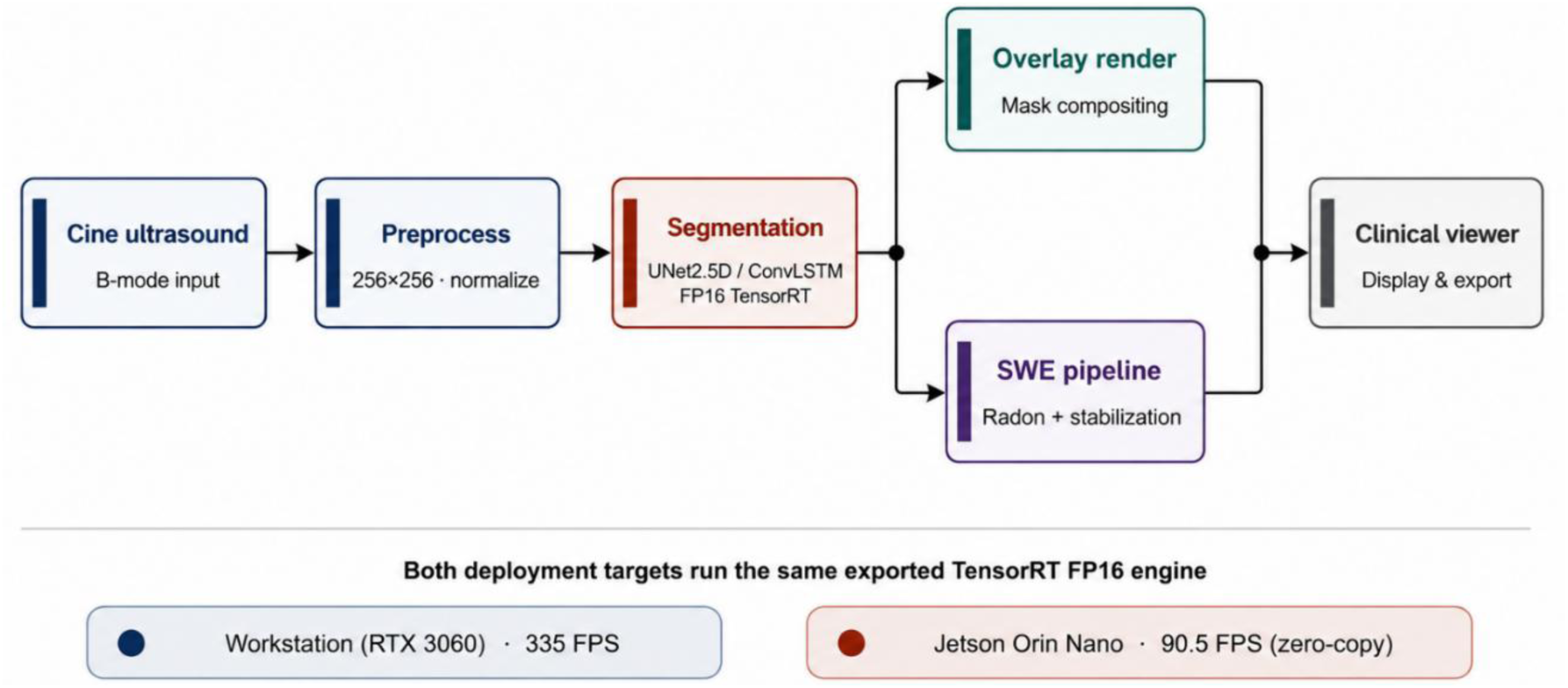
MyoStat.AI pipeline overview. From cine ultrasound input through preprocessing, segmentation (UNet2.5D / ConvLSTM, FP16 TensorRT), the parallel overlay-render and SWE branches, and final display in the clinical viewer. Each deployment target runs a TensorRT engine compiled from the same exported ONNX model.

### Segmentation Accuracy

The segmentation accuracy of the proposed framework is evaluated across different model architectures and deployment scenarios, demonstrating both high fidelity and efficient execution. The architectures were evaluated using Dice, Intersection over Union (IoU), and latency across the same conditions (**Table 7**, **Figure 2**). The ConvLSTM model achieved the highest scores among the tested architectures, with a Dice coefficient of 0.994 and an IoU of 0.987 on the training and validation sets. This is an absolute increase of 18.6 percentage points over the 2D baseline (Dice 0.808, IoU 0.678). UNet2.5D and UNet3D achieved intermediate Dice coefficients of 0.983 and 0.988, and IoU scores of 0.967 and 0.977, respectively. These differences reflect single-run results under the tested ablation configuration; no repeated-run statistics or hypothesis testing were performed, so the observed ranking should be read as descriptive rather than as evidence of statistically significant superiority, particularly given the small margin between UNet2.5D, UNet3D, and ConvLSTM.

**Figure 2.**
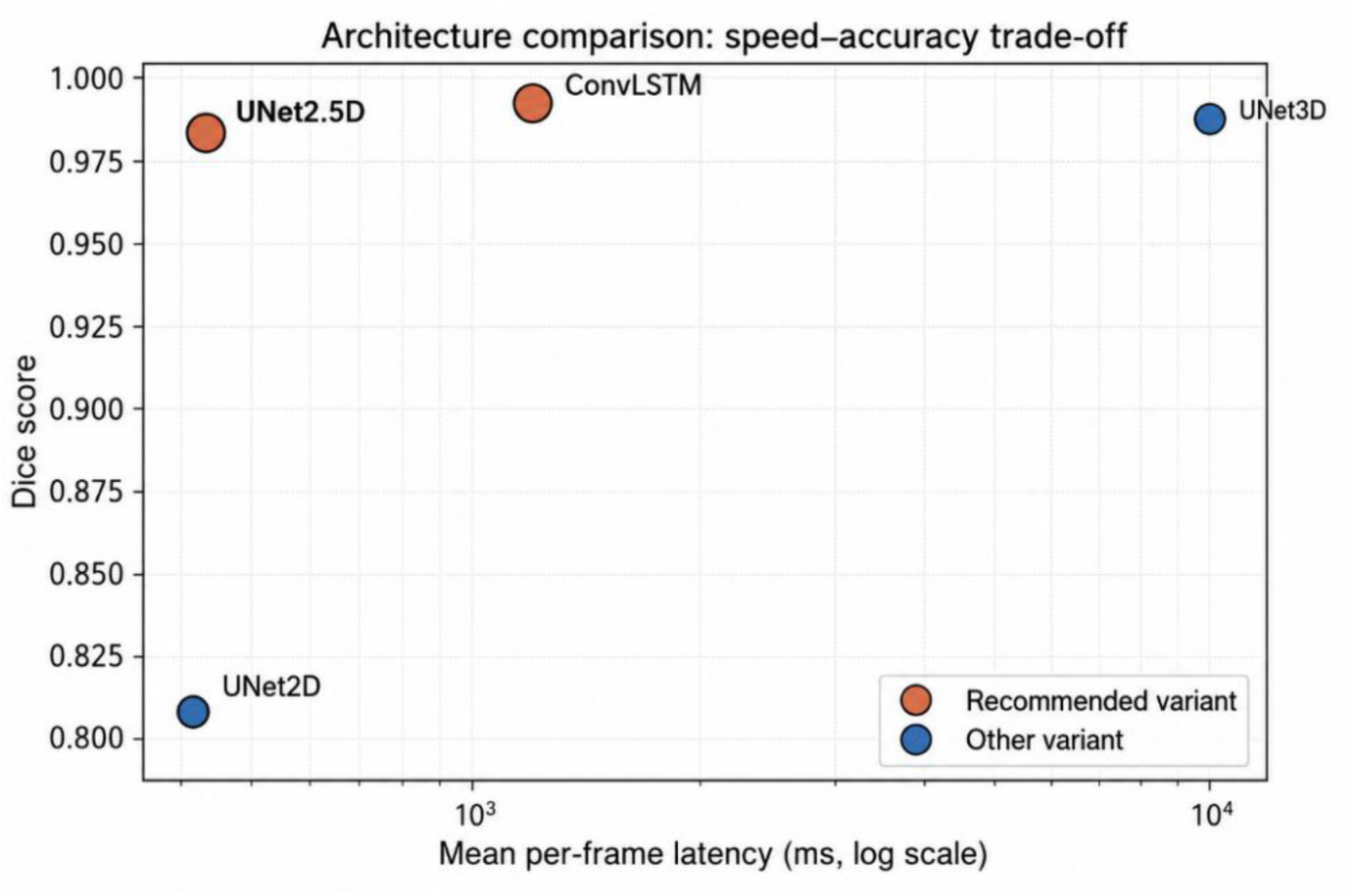
Accuracy–latency trade-off across segmentation architectures. Comparison across UNet2D, UNet2.5D, ConvLSTM, and UNet3D. Dice score plotted against mean per-frame inference latency on a log scale. ConvLSTM and UNet2.5D are highlighted as the accuracy-priority and latency-conscious recommendations.

**Table 7.** Segmentation Accuracy & Latency Comparisons.

| Metric | UNet2D baseline | UNet2.5D | UNet3D | UNet + ConvLSTM |
| --- | --- | --- | --- | --- |
| Dice | 0.808 | 0.983 | 0.988 | 0.994 |
| IoU | 0.678 | 0.967 | 0.977 | 0.987 |
| Mean latency (ms) | 417 | 433 | 9976 | 1193 |
| Latency standard deviation (ms) | 12 | 25 | 3608 | 99 |
| p95 latency (ms) | 434 | 466 | 14735 | 1370 |
| Parameters (M) | 7.7 | 7.7 | 22.26 | 9.36 |

The computational efficiency of the models was further assessed by benchmarking their inference latency. The ConvLSTM model demonstrated a mean per-frame latency of ∼1193 ms, while the 2.5D approach exhibited a mean per-frame latency of ∼433 ms, the baseline UNet2D ∼417 ms, and the UNet3D architecture ∼9976 ms. Additional hyperparameters such as temporal window size, loss function, augmentation strategy, and dropout rate were also varied on the UNet2.5D backbone against the Dice score. The benchmark results identified the highest-Dice configuration in this ablation as a temporal window size of 3 frames, the BCE loss function, full augmentation strategy, and a dropout rate of 0.2. Each value is the best-scoring point along its own single-variable axis rather than a jointly trained configuration; no single model was trained with all four settings combined. The UNet2.5D model was then exported to an FP16 TensorRT engine for deployment (NVIDIA, 2024b). The deployed UNet2.5D engine, which is the model benchmarked in **Figure 3**, achieved 341 FPS on the RTX 3060 and 90.4 FPS on the Jetson Orin Nano. ConvLSTM’s higher accuracy carries a much higher per-frame cost (1193 ms on CPU; **Table 7**) and it was not the model used for the reported edge throughput.

**Figure 3.**
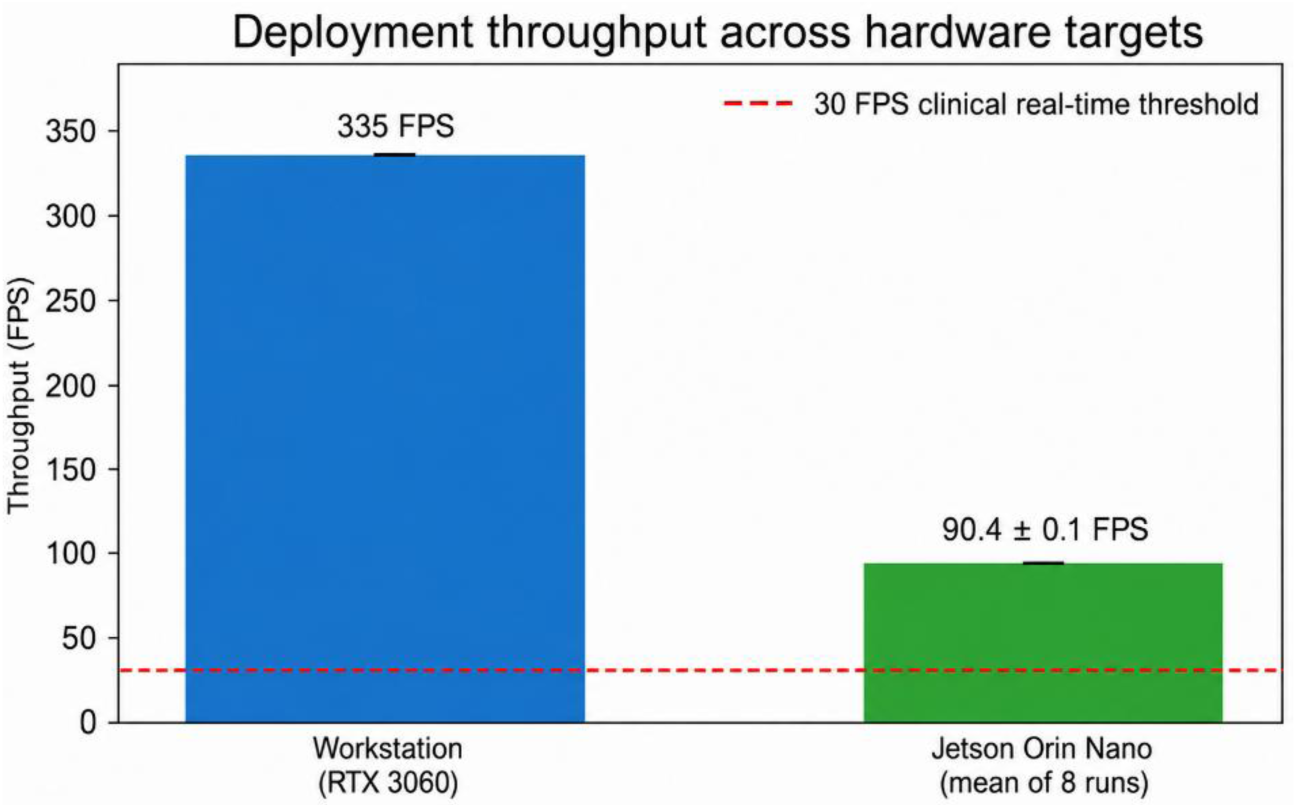
Deployment throughput across hardware targets. UNet2.5D FP16 TensorRT throughput on the workstation (RTX 3060, 341 FPS) and on the Jetson Orin Nano (mean 90.4 FPS across 8 runs), with the 30 FPS real-time inference target (defined in Section 2.1.5) marked.

### Evaluation of SWE Stability

The temporal stability and robustness of the estimated shear-wave velocity fields, not the segmentation masks, were assessed by evaluating the impact of six distinct stabilization methods applied to the temporal sequence, with the primary evaluation metric employed being Temporal Coefficient of Variation (CoV), which measures the frame-to-frame consistency of the estimated velocity field. Three main categories of stabilization were compared: no stabilization (baseline), smoothn presets, and SNR-adaptive methods.

All three penalized least-squares variants reduced the baseline CoV from 81.07 to 29.37, a 63.8% improvement, with no meaningful difference between λ values (**Table 8**). This saturation indicates that smoothn reaches its effective ceiling on this synthetic corpus at λ=5; increasing the penalty further provides no additional stability gain. Among the adaptive methods, adaptive_coarse achieved the strongest raw reduction, from 81.07 to 6.04 (92.6%), but at the cost of severe signal distortion. Mean velocity dropped from 3.11 to 0.90 m/s and edge preservation collapsed to 0.317, indicating that the aggressive window sizes over-smooth the velocity field and distort the underlying SWE signal. adaptive_fine performed inversely, increasing CoV from 81.07 to 130.67 (−61.2%), destabilizing rather than smoothing the velocity field on this input.

**Table 8.** SWE Stability & Latency Results.

| Method | CoV | CoV Improvement | Latency Overhead | Edge Preservation | Pass? |
| --- | --- | --- | --- | --- | --- |
| **None (baseline)** | **81.07** | **—** | **2.1 ms** | **1.000** | **No** |
| smoothn conservative $\lambda=5$ | 29.37 | 63.8% | 5.0 ms | 1.000 | Yes |
| smoothn moderate $\lambda=10$ | 29.37 | 63.8% | 2.7 ms | 1.000 | Yes |
| smoothn aggressive $\lambda=20$ | 29.37 | 63.8% | 2.0 ms | 1.000 | Yes |
| adaptive_fine | 130.67 | -61.2% | 3.4 ms | 0.421 | Yes |
| adaptive_coarse | 6.04 | 92.6% | 0.9 ms | 0.317 | Yes |

A subsequent analysis was also performed to investigate the computational cost of each stabilization method. All six methods remained well within the pre-defined 20ms latency overhead threshold. The smoothn variants introduced overheads of 5.0ms (conservative), 2.7ms (moderate), and 2.0ms (aggressive), while the adaptive methods introduced 3.4ms (adaptive_fine) and 0.9ms (adaptive_coarse). Latency is therefore not a differentiating factor between methods at this data scale. These figures are per-frame computed overhead only. Because the stabilization window spans 25 frames at 30 Hz (**Table 3**), a causal real-time implementation would additionally incur roughly 0.83 s of window and buffering delay before a stabilized estimate is available, and this delay (not the millisecond-scale compute overhead) governs end-to-end responsiveness. All pass the acceptance criterion, and the preset recommendation is determined entirely by stability and signal fidelity.

The Python implementation follows the smoothn algorithm of Garcia (2010) directly. Numerical equivalence between the Python and the original MATLAB implementation was assessed via Pearson correlation on matched outputs; full MAE and RMSE comparison against a MATLAB reference is planned as a validation step prior to clinical deployment. Under the tested synthetic conditions, the smoothn_moderate preset (λ=10) is provisionally recommended as a starting configuration for future real-time evaluation, pending validation on real acquisitions. It achieved 63.8% CoV reduction at 2.7ms of latency overhead per frame while maintaining an edge-preservation score of 1.000 on this synthetic corpus. Although smoothn_aggressive offered marginally lower overhead (2.0ms), it delivered the same CoV reduction. The moderate setting is preferred as the more conservative choice for future testing on real clinical data, where the cost of over-smoothing tissue boundaries is expected to be higher than on the synthetic velocity fields used here; this preference is a hypothesis to be tested, not a clinical recommendation, and no preset should be adopted clinically prior to validation against real acquisitions. **Reproducibility Verification & Failure Mode Analysis.** To assess the repeatability of inference timing, a benchmark harness was implemented based on the methodology described in methods section. This harness verifies deterministic execution and data integrity across independent runs of the same trained model on the same hardware and software stack. Prior to each benchmark run, SHA-256 checksums were verified against the dataset manifest to ensure data integrity. All 8 runs passed manifest verification with matching checksums, confirming no silent data drift occurred between runs. Because each run times the same fixed TensorRT engine on identical hardware, latency repeatability does not depend on a random seed. Each run produced a timestamped evidence bundle containing configuration files, per-run metrics, and a manifest.json with SHA-256 checksums of all output files, with a Git commit hash (27404e3a) recorded at generation time for traceability. This procedure measures whether repeated executions of an already-trained, fixed configuration yield consistent timing, it does not assess whether model training itself, or the resulting accuracy metrics, are reproducible across different initializations.

The multi-run timing analysis reported a cross-run coefficient of variation of 0.174% (**Figure 4**), a mean inference latency of 11.062 ms (±0.019 ms cross-run σ), and a latency range across runs of 0.055 ms. The mean within-run standard deviation was 0.044 ms, and mean throughput was 90.4 FPS. All runs maintained p95 latency below 11.13 ms and sustained ≥90 FPS on the Jetson (341 FPS on the workstation), comfortably exceeding the 30 FPS real-time engineering inference target defined in the methods section.

**Figure 4.**
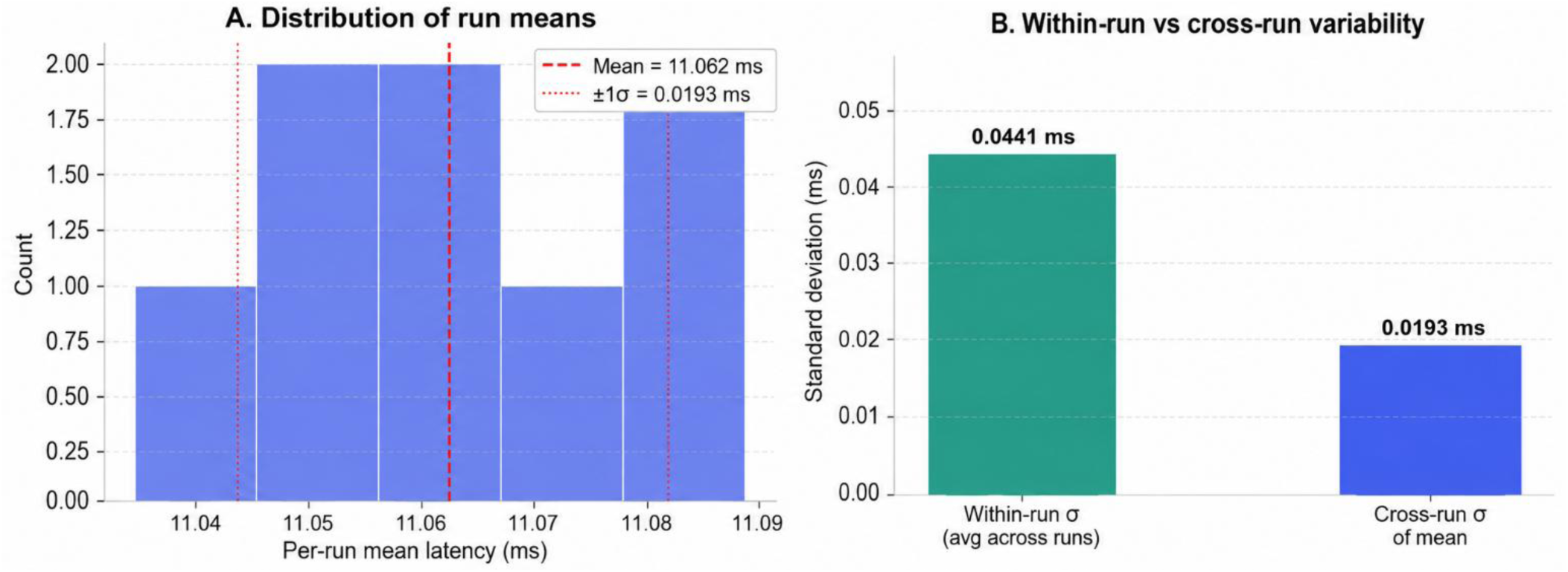
Reproducibility of inference-latency benchmarking on the Jetson Orin Nano. Eight independent benchmarking runs were performed to assess both within-run and between-run variability in per-frame inference latency. (**A**) Distribution of the mean latency calculated for each run. The red dashed line indicates the overall mean latency across all runs (11.062 ms), while the red dotted lines denote ±1 standard deviation of the run means (±0.0193 ms). The narrow distribution of run-level means corresponds to a coefficient of variation of 0.174%, demonstrating highly consistent performance across independent executions. (**B**) Comparison of the average within-run standard deviation and the cross-run standard deviation of the mean latency. The mean within-run variability was 0.0441 ms, whereas the variability between run means was substantially lower at 0.0193 ms. These results indicate that short-term frame-to-frame fluctuations within individual runs were small and that the deployed inference pipeline maintained highly reproducible latency across repeated benchmark sessions.

The low cross-run CoV indicates that, for this fixed model, engine, and hardware configuration, inference timing is highly consistent across repeated executions. This is evidence of timing repeatability only, it does not validate the deterministic configuration protocol’s effect on training outcomes, nor does it establish that reported accuracy metrics (Dice, IoU, CoV) are reproducible across independent training runs. The approximately 2.3:1 ratio of within-run to cross-run standard deviation (corresponding to roughly a 5.2:1 variance ratio) suggests that, for this specific configuration, system-level timing non-determinism contributes less to the observed latency distribution than frame-content-dependent dynamics, though this has not been tested as a designed analysis. Whether single-run benchmarks with sufficient warmup are representative of multi-run timing behavior for other hardware-model configurations has not been evaluated.

The current analysis carries limitations more consequential than timing repeatability. Training and evaluation masks are produced by the same synthetic generator, so the reported Dice, IoU, and CoV values reflect internal consistency with the generator’s own geometry rather than independently verified anatomical accuracy, an evaluation setup that cannot by itself demonstrate the pipeline recognizes real myocardial boundaries, speckle patterns, or motion. No clinical labels, patient identifiers, or real cardiac acquisitions were used at any stage, so tissue heterogeneity, operator variability, and acquisition artifacts intrinsic to real echocardiography remain entirely unassessed. Secondary, to these domain limitations, all runs also used a single serialized engine on a single hardware unit, so neither timing nor accuracy has been evaluated for sensitivity to initialization, hardware, or site. Multi-seed training reproducibility and multi-site timing validation are recommended future work but resolving them would not address the more fundamental synthetic-domain and validation gap described above. The performance metrics of the segmentation process are closely linked to the failure modes identified previously (**Figure 5**). Analyzing the impacts of these failure modes is critical for understanding the specific conditions under which the model exhibits performance degradation and produces unreliable outputs.

**Figure 5.**
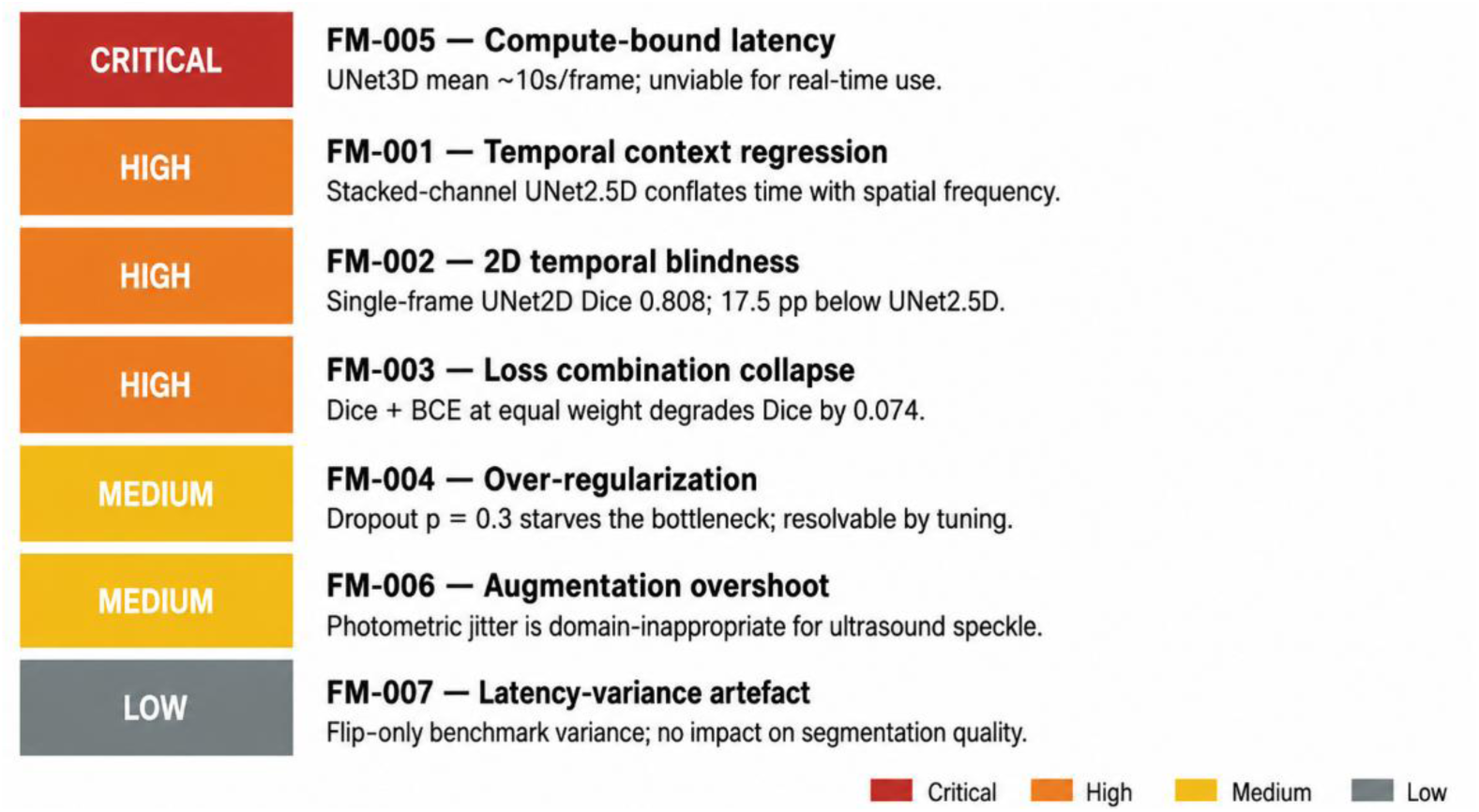
Engineering severity tiers across the seven failure modes. Failure mode summary across the seven characterized modes, grouped by post hoc operational severity (not a validated clinical classification). Critical: latency or accuracy makes the tested configuration unusable for real-time inference regardless of other factors. High: accuracy degradation exceeding 15 percentage points from the best-performing variant on the same axis. Medium: resolvable through hyperparameter changes alone, with degradation under 15 points. Low: a benchmarking artifact with no measured accuracy impact.

Temporal context regression (FM-001) is classified as high severity. It is caused when the stacked-channel approach in UNet25D treats frames as spatial channels, conflating temporal displacement with spatial frequency. Performance peaks at a 3-frame window (Dice=0.984, +6.4pp over single-frame) and decreases monotonically at 5 frames (Dice=0.976) and 7 frames (Dice=0.956), indicating that the stacked-channel representation degrades as the temporal span extends beyond the immediate neighboring frames (**Table 9**). Mitigation strategies for this failure mode include using the ConvLSTM bottleneck, adding temporal positional encoding to distinguish frame order from spatial content, pre-training the model on 1-frame and fine-tuning on multi-frame data, or weighting the central frame higher to reduce sensitivity to window length.

**Table 9.** Segmentation Accuracy Metrics across Temporal Frame Sizes.

| Variant | Dice | IoU | $\Delta$ Dice vs 1-frame |
| --- | --- | --- | --- |
| 1-frame (baseline) | 0.920 | 0.852 | — |
| 3-frame | 0.984 | 0.969 | +0.064 |
| 5-frame | 0.976 | 0.954 | +0.057 |
| 7-frame | 0.956 | 0.916 | +0.037 |

Temporal Blindness (FM-002) is classified as high severity and is caused when only single ultrasound frames are provided. This may be insufficient at certain cardiac phases; one hypothesis is that temporal models implicitly exploit frame-to-frame boundary consistency as a regularizing signal, but this mechanism has not been directly tested and should not be treated as established (**Table 10**). Mitigation strategies include stacking 3 frames as channels to inject temporal context at negligible parameter cost, adding anatomical shape priors (e.g. level-set or ellipse prior loss) to constrain 2D predictions, or utilizing test-time augmentation (TTA) with temporal jitter to probe boundary stability.

**Table 10.** Segmentation Accuracy Metric Comparison for Single Frame Inputs.

| Variants | Dice | IoU | Params (M) | Latency mean (ms) |
| --- | --- | --- | --- | --- |
| UNet2D (baseline) | 0.808 | 0.679 | 7.70 | 402 |
| UNet25D | 0.983 | 0.967 | 7.70 | 422 |
| ConvLSTM | 0.994 | 0.987 | 9.36 | 1123 |
| UNet3D | 0.988 | 0.977 | 22.26 | 9715 |

Loss Function Step-Budget Sensitivity (FM-003) is classified as high severity. Under a limited training step budget, pure Dice loss is the weakest performer (Dice=0.917) due to gradient sparsity early in training, where the global overlap objective provides weak signal when activations are near uniform. BCE alone achieves the highest score in this axis (Dice=0.984), while combined losses are intermediate: Dice+BCE (0.967) and Dice+Focal (0.947). The results indicate sensitivity to step budget rather than a categorical failure of any specific loss combination (**Table 11**). Mitigation strategies include using alternate loss functions or combinations, applying gradient normalization to balance multi-term losses dynamically, and using a learning-rate warm-up schedule when training with combined losses to prevent early instability.

**Table 11.**
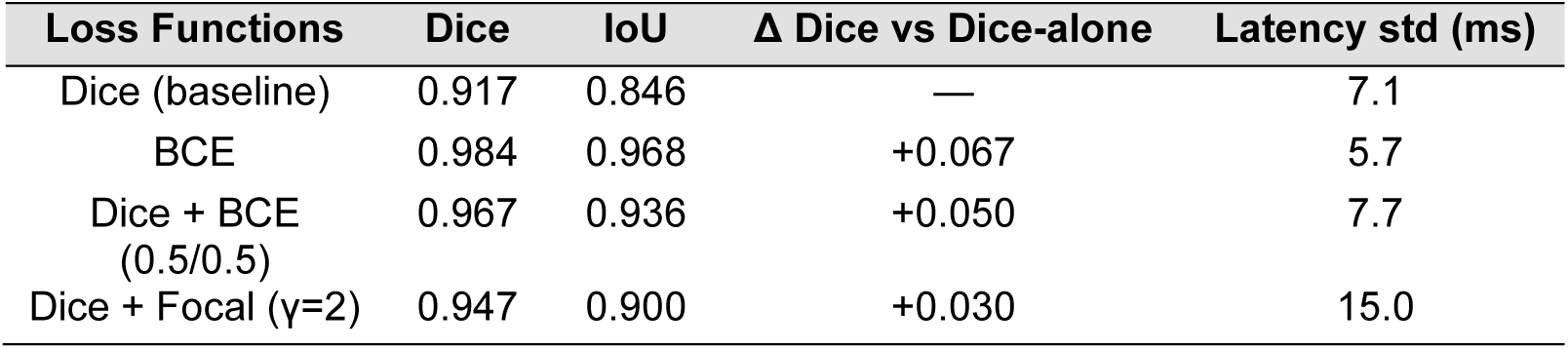
Segmentation Accuracy Metric across Loss Functions.

| Loss Functions | Dice | IoU | $\Delta$ Dice vs Dice-alone | Latency std (ms) |
| --- | --- | --- | --- | --- |
| Dice (baseline) | 0.917 | 0.846 | — | 7.1 |
| BCE | 0.984 | 0.968 | +0.067 | 5.7 |
| Dice + BCE<br>(0.5/0.5) | 0.967 | 0.936 | +0.050 | 7.7 |
| Dice + Focal ($\gamma=2$) | 0.947 | 0.900 | +0.030 | 15.0 |

Over-Regularization (FM-004) is classified as medium severity. The dropout axis produces a non-monotonic response: p=0.1 is the worst performer (Dice=0.909, below the no-dropout baseline of 0.923), while p=0.2 achieves the best result (Dice=0.967) and p=0.3 is essentially tied (Dice=0.966) (**Table 12**). The failure at p=0.1 is consistent when dropout is sufficient to disrupt gradient flow but insufficient to force the weight redundancy that makes higher rates beneficial. Thus, p=0.2 is the recommended default. Mitigation strategies include using p=0.2 as the recommended default, applied to the bottleneck only to preserve encoder and decoder spatial features. p=0.1 should be avoided as it disrupts gradient flow without providing the weight redundancy that makes higher rates beneficial. If a higher degree of regularization is required, p=0.3 is an acceptable alternative given its near-identical performance to p=0.2.

**Table 12.**
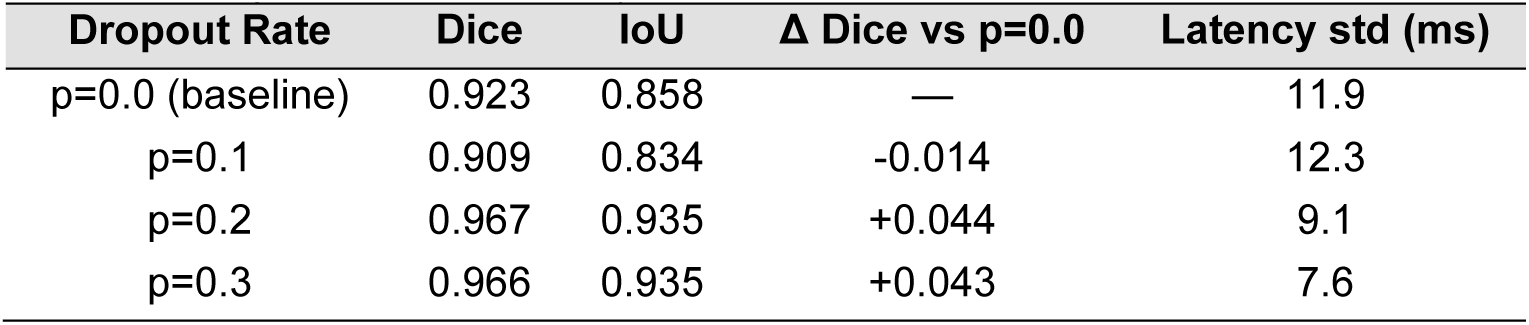
Segmentation Accuracy Metric across Dropout Rates.

Compute-Bound Latency (FM-005) is classified as critical severity. The UNet3D model achieves competitive segmentation (Dice=0.988) but is 24 times slower than UNet25D at mean latency 9976ms, with p95=14735ms. This renders it unviable for real-time clinical use. Mitigation strategies include deploying the UNet25D or ConvLSTM models instead, reducing UNet3D depth (fewer encoder stages) or using separable 3D convolutions (depthwise + pointwise) to cut FLOPs, or training a smaller student model on UNet3D soft labels.

Augmentation Overshoot (FM-006) is classified as medium severity. Flip+Rotation is the sole augmentation configuration that degrades accuracy below the no-augmentation baseline (Dice=0.873, Δ−0.079 vs aug_none). Adding rotation without complementary photometric augmentation shifts the training distribution in a way that does not match inference-time frames. Full augmentation, including brightness/contrast jitter, is the strongest performer in this axis (Dice=0.972, Δ+0.020), suggesting that photometric jitter compensates for the geometric shift introduced by rotation rather than compounding it (**Table 13**). Mitigation strategies include avoiding flip+rotation as a standalone configuration. Either flip-only or the full augmentation pipeline (flip, rotation, brightness/contrast jitter) are preferable. Ultrasound-specific augmentations (speckle noise injection, acoustic shadow simulation) should be evaluated as additional strategies once real acquisition data becomes available.

**Table 13.**
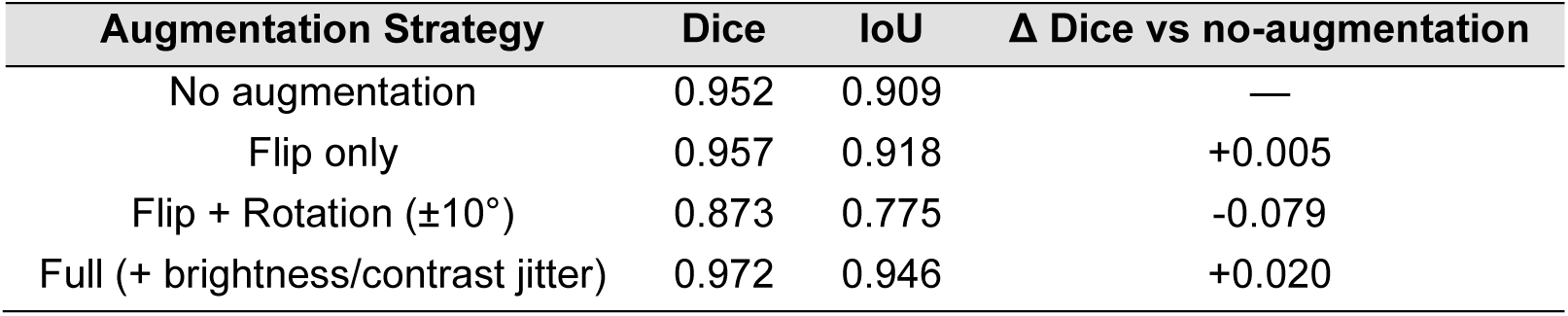
Segmentation Accuracy Metric across Augmentation Strategies.

| Augmentation Strategy | Dice | IoU | $\Delta$ Dice vs no-augmentation |
| --- | --- | --- | --- |
| No augmentation | 0.952 | 0.909 | — |
| Flip only | 0.957 | 0.918 | +0.005 |
| Flip + Rotation ($\pm 10^\circ$) | 0.873 | 0.775 | -0.079 |
| Full (+ brightness/contrast jitter) | 0.972 | 0.946 | +0.020 |

Latency Variance Across Separately Trained Augmentation Variants (FM-007) is classified as low severity. Canonical benchmarking on the workstation CPU (PyTorch, FP32; the same pipeline used for **Table 9-13**) shows tightly clustered latency across all four augmentation variants (mean 929–941 ms, p95/mean 1.01–1.03), with low variance (**Table 14**). The flip-only result reported in earlier analysis (mean = 690 ms, std = 506 ms, p95 = 1533 ms) was not reproducible in canonical testing and is attributed to an environment-specific scheduling event rather than a property of the augmentation pipeline. Because inference-time computation is identical regardless of which augmentation was used during training, this latency variation reflects run-to-run measurement noise across separately trained model instances rather than an effect of augmentation on inference. Segmentation accuracy across these same variants ranges from Dice 0.873 to 0.972 (**Table 13**) and is reported and discussed there; it is not a finding of this latency analysis. As no anomalous latency variance was observed in canonical testing, no mitigation is required at this time. If variance above the 1.5× p95/mean threshold is observed in future hardware configurations, isolated CPU affinity and median-based latency reporting are recommended diagnostic steps.

**Table 14.** Processing Latency across Augmentation Strategies.

| Augmentation Strategy | Latency mean (ms) | Latency std (ms) | Latency p95 (ms) | p95/mean |
| --- | --- | --- | --- | --- |
| No augmentation | 941 | 15.9 | 967 | 1.03 |
| Flip only | 931 | 13.4 | 959 | 1.03 |
| Flip + Rotation ($\pm 10^\circ$) | 932 | 11.1 | 953 | 1.02 |
| Full | 929 | 5.9 | 940 | 1.01 |

## Discussion

### Performance vs Prior work

The significance of these results becomes more apparent when comparing previous works. Notably, the work done by Ronneberger et al. reported the UNet model achieving an IoU score of 0.92 and 0.78 for the PhC-U373 and DIC-HeLa datasets which outperformed the best models by a large margin back in 2015 (Ronneberger et al., 2015). This work demonstrated the potential of UNet architecture for complex medical image segmentation. Further studies also show the effectiveness of the UNet architecture. One such study was by Leclerc, where the performance of the U-Net architecture was evaluated on the CAMUS dataset and reported a Dice score of ∼0.94 (Leclerc et al., 2019). CAMUS is a clinically acquired dataset spanning good-, medium-, and poor-quality image subgroups, not a good/high-quality-only selection. Our results indicate a lower Dice score of 0.808 for the baseline UNet2D model (**Table 7);** because our evaluation uses synthetic data while Leclerc et al. use real clinical acquisitions under a different architecture, training configuration, and preprocessing pipeline, this gap cannot be attributed to any single factor without a matched experiment on the same dataset and settings. The comparison is reported here for context only and should not be read as explaining or excusing the difference in baseline performance. This comparison highlights the improvement achieved by ConvLSTM architecture. This sequence-aware architecture allows our model to leverage contextual information, achieving the highest Dice score under the tested synthetic conditions, though at a substantially higher CPU latency (1193 ms mean; **Table 7**) than the 2D and 2.5D variants.

### Speed–Accuracy Trade-off and Architecture Choice

Hyperparameter tuning showed the UNet2.5D backbone is sensitive to configuration choices, with augmentation strategy producing the largest swing in this axis (Dice 0.873 for flip+rotation alone vs. 0.972 for full augmentation, a range of 0.099), followed by dropout rate (Dice 0.909 at p=0.1 vs. 0.967 at p=0.2, Δ +0.058) and loss function (Dice 0.917 for Dice-alone vs. 0.984 for BCE-alone, Δ +0.067). Temporal window size showed a smaller but still material effect, peaking at a 3-frame window (Dice 0.984, Δ +0.064 vs. single-frame) and declining at longer windows (**Table 9**). When taking the total latency into account, the UNet 2D and UNet 2.5D architectures are significantly faster than the ConvLSTM, but these two architectures fall short when looking at the accuracy comparisons between the ConvLSTM and UNet 3D models (**Table 7**, **Figure 2**). The outcome suggests that the ConvLSTM model serves as a tolerable middle ground achieving accuracy results on par with the UNet 3D model while balancing it with manageable computational demands, offering a valuable trade-off between performance and efficiency that is superior to the established methods under the tested synthetic conditions.

### Limitations and Synthetic-data Scope

The result of our experiment runs indicate that our optimized models sustained real-time inference throughput across diverse hardware environments, achieving 341 FPS on a high-performance GPU (RTX 3060) and 90.4 FPS on a resource-constrained embedded platform (Jetson Orin Nano) (**Figure 3**). However, while the current results establish high accuracy metrics on the training and validation sets, it is important to acknowledge the limitations inherent in this scope. These findings are confined to the specific datasets utilized in this research, meaning they do not yet represent external validation of the model’s generalization capability. Therefore, these results serve as a strong foundation for the proposed method but must be interpreted cautiously regarding their applicability to entirely novel, unseen data. The temporal Coefficient of Variation is used as the primary stability metric because it quantifies frame-to-frame temporal variability of the SWS estimates. CoV measures variability only and must be interpreted alongside accuracy and fidelity, since a low CoV can coexist with a biased or over-smoothed estimate, as adaptive_coarse demonstrates: a high CoV indicates that velocity values fluctuate substantially across frames, producing an elastography map that would be difficult to interpret or reproduce consistently. Reducing CoV through post-processing stabilization therefore yields more temporally uniform velocity-field under the tested synthetic conditions. Validation against real acquisitions is still required, however, to determine whether this translates into clinically worthy SWE measurements.

A key limitation of these results is that all stability evaluations were performed on synthetic velocity fields generated from a fixed random seed, not physical phantoms or real acquisitions. The term “phantom” is reserved here for a physical or simulated test object with known imaging properties, which the current velocity-field generator does not represent. Real cardiac SWE data introduces confounds absent from synthetic corpora: tissue heterogeneity across myocardial layers, respiratory motion, probe-pressure variation, and acoustic clutter from rib shadows and near-field reverberation. The CoV reductions reported here should therefore be interpreted as method-to-method comparisons under controlled conditions rather than absolute predictions of in vivo performance. Validation against real acquisitions is the necessary next step before any preset is adopted clinically.

### Failure mode analysis

The severity levels used throughout this analysis are operational engineering categories, not clinically validated classifications. Critical denotes modes that make a configuration unviable for real-time use regardless of accuracy; high denotes large accuracy degradation with a plausible architectural cause; medium denotes modes addressable through hyperparameter changes alone; low denotes a benchmarking artifact with no accuracy impact. No clinician input or prespecified clinical requirement informed these tiers. Two patterns are reasonably well supported by the ablation data. Temporal context helps, and how it is encoded matters: single-frame input underperforms every multi-frame variant (FM-002), and UNet2.5D’s stacked-channel encoding shows a non-monotonic response that degrades beyond a 3-frame window (FM-001; **Tables 9–10**). ConvLSTM performed best on both axes under the tested conditions, though whether this holds for longer sequences, other cardiac phases, or real data remains untested. Separately, UNet3D’s CPU FP32 latency and variance (FM-005; **Table 7**: mean 9976 ms, p95 14,735 ms, approximately 23× UNet2.5D) are high enough to be impractical for real-time use as currently implemented. No GPU-optimized or TensorRT UNet3D configuration was benchmarked, so this conclusion is scoped to the tested CPU implementation and is not a categorical rejection of the architecture.

Beyond these two points, the current data support an observed ranking, not a mechanism. The loss-function, dropout, and augmentation results (FM-003, FM-004, FM-006) each come from a single 20-gradient-step run per variant with per-variant rather than matched seeding, so a genuine effect cannot yet be distinguished from run-to-run variance. The mechanistic explanations related to gradient sparsity, weight redundancy, distributional shift, are hypotheses the current experiment cannot support and should not be treated as established causes. FM-007’s latency clustering across augmentation variants is a workstation CPU FP32 benchmarking artifact, distinct from the TensorRT FP16 Jetson deployment timings, and the two figures should not be compared directly.

Taken together, these findings motivate a two-stage validation plan rather than a final configuration. Architectural choices, temporal encoding and 3D compute cost, warrant re-testing first, since they show the largest and most consistent effects; hyperparameter choices should be re-run with matched seeds and multiple repeats before any default is fixed, since the current single-run rankings are not distinguishable from noise. Both stages require converged training and, eventually, evaluation against real acquisition data before any configuration is treated as preferred.

## Conclusions

MyoSTAT.AI demonstrates a deterministic, reproducible cardiac segmentation and SWE benchmarking framework, with the ConvLSTM variant achieving Dice 0.994 and smoothn-based stabilization yielding a 63.8% CoV reduction. While the UNet2.5D variant being able to achieve a throughput of 90.4 FPS on Jetson Orin Nano via TensorRT FP16 making it promising for resource-constrained edge computing platforms. These results are based entirely on synthetic data; validation on real clinical acquisitions, along with resolution of acquisition-compatibility and cross-model numerical discrepancies noted in this work, remains necessary before any deployment claim can be made.

## Code, Data, and Materials Availability

Repository link and DOI and evidence bundle available upon request and review of authors.

## Data Availability

All data produced in the present study are available upon reasonable request to the authors

## Acknowledgments

This research received no specific grant from any funding agency in the public, commercial, or not-for-profit sectors. The authors acknowledge institutional and computational support from the University of Toronto Mississauga, the University of Toronto Department of Medical Biophysics, Yale University Department of Therapeutic Radiology, and Northeastern University Toronto Department of Biotechnology.

